# Management and containment of a SARS-CoV-2 Beta variant outbreak at the Malawi University of Science and Technology

**DOI:** 10.1101/2021.11.25.21266298

**Authors:** Gama Bandawe, Petros Chigwechokha, Precious Kunyenje, Yohane Kazembe, Jeverson Mwale, Madalitso Kamaliza, Mtisunge Mpakati, Yanjanani Nyakanyaka, Charles Makamo, Saizi Kimu, Mwayiwawo Madanitsa, Joseph Bitilinyu Bangoh, Tonney Nyirenda, Richard Luhanga, Martha Sambani, Benard Mvula, Jennifer Giandhari, Sureshnee Pillay, Yeshnee Naidoo, Upasana Ramphal, James Emmanuel San, Houriiyah Tegally, Eduan Wilkinson, Tulio de Oliveira, Address Malata

## Abstract

Outbreaks of COVID at university campuses can spread rapidly and threaten the broader community. We describe the management of an outbreak at a Malawian university in April-May 2021 during Malawi’s second wave. Classes were suspended following detection of infections by routine testing and campus-wide PCR mass testing was conducted. Fifty seven cases were recorded, 55 among students, two among staff. Classes resumed 28 days after suspension following two weeks without cases. Just 6.3% of full-time staff and 87.4% of outsourced staff tested while 65% of students at the main campus and 74% at the extension campus were tested. Final year students had significantly higher positivity and lower testing coverage compared to freshmen. All viruses sequenced were beta variant and at least four separate virus introductions onto campus were observed. These findings are useful for development of campus outbreak responses and indicate the need to emphasize staff, males and senior students in testing.

**Article Summary Line:** Successful management of a campus outbreak using test trace and isolate approach with resumption within a month following suspension of all in-person classes. Trends in voluntary testing by gender, age and year of study that can help in formation of future management approaches.

## Introduction

Educational institutions such as boarding schools and universities are considered important focal points in the transmission of SARS-CoV-2 throughout the broader community (1).Once transmission is established in a semi-closed university community, growth of the epidemic is expected to be rapid and control becomes extremely difficult (2). With outbreaks observed at university campuses globally (3) and in schools around Malawi, local authorities under the guidance of the Presidential Task Force on COVID-19 ordered the closure of all educational institutions from March to October 2020 (4) (5). During Malawi’s second wave of infections towards the end of 2020 and early 2021 and following an outbreak in a school in Lilongwe where 311 out of 605 students tested positive, schools were again closed for three weeks from 18^th^ January 2021 (5).

At the Malawi University of Science and Technology (MUST) in-person classes resumed in March with a phased and cautious blended learning approach. This included online learning for all students and face-to-face classes with only 50% of students returning to campus in two staggered groups. Other measures implemented included enforcement of COVID-19 prevention measures such as mask wearing, hand washing and physical distancing. The MUST SARS-CoV-2 testing laboratory was also set up to conduct RT-PCR and rapid antigen testing of all individuals presenting at MUST Clinic with any COVID-19 related symptoms.

Despite all these measures residence halls and campus lecture rooms remained susceptible to outbreaks due to the continued movement of students in and out of campus especially to Blantyre, Malawi’s second largest city 30 kilometres away where 38% of the average daily national cases for the month of March were located.

During the months of April and May, 2021 MUST experienced an outbreak of COVID-19 involving 57 confirmed cases in a population of 1152 (∼5%) students. In line with Ministry of Health guidelines, students were not sent home but kept on campus and a containment strategy of enhanced testing capacity accompanied by contact tracing and isolation (TTI) of known cases (6,7) was used.

Here, we describe the outbreak and the management strategy that enabled the resumption of in-person classes 28 days after identification of the first case. By analysing administrative data from the outbreak, we identify trends and patterns, which emerged from implementation of the response plan.

These experiences, lessons and findings may prove useful to other educational institutions and decision-makers in the planning and development of responses to future campus outbreaks especially in the African context where similar challenges may exist.

## Materials and methods

### MUST setting and demographics

The Malawi University of Science and Technology is a public university situated at Ndata Estate in Thyolo district in southern Malawi (Figure 1). It is in a rural setting approximately 30 kilometres from Blantyre, Malawi’s second largest city.

**Figure 1:**
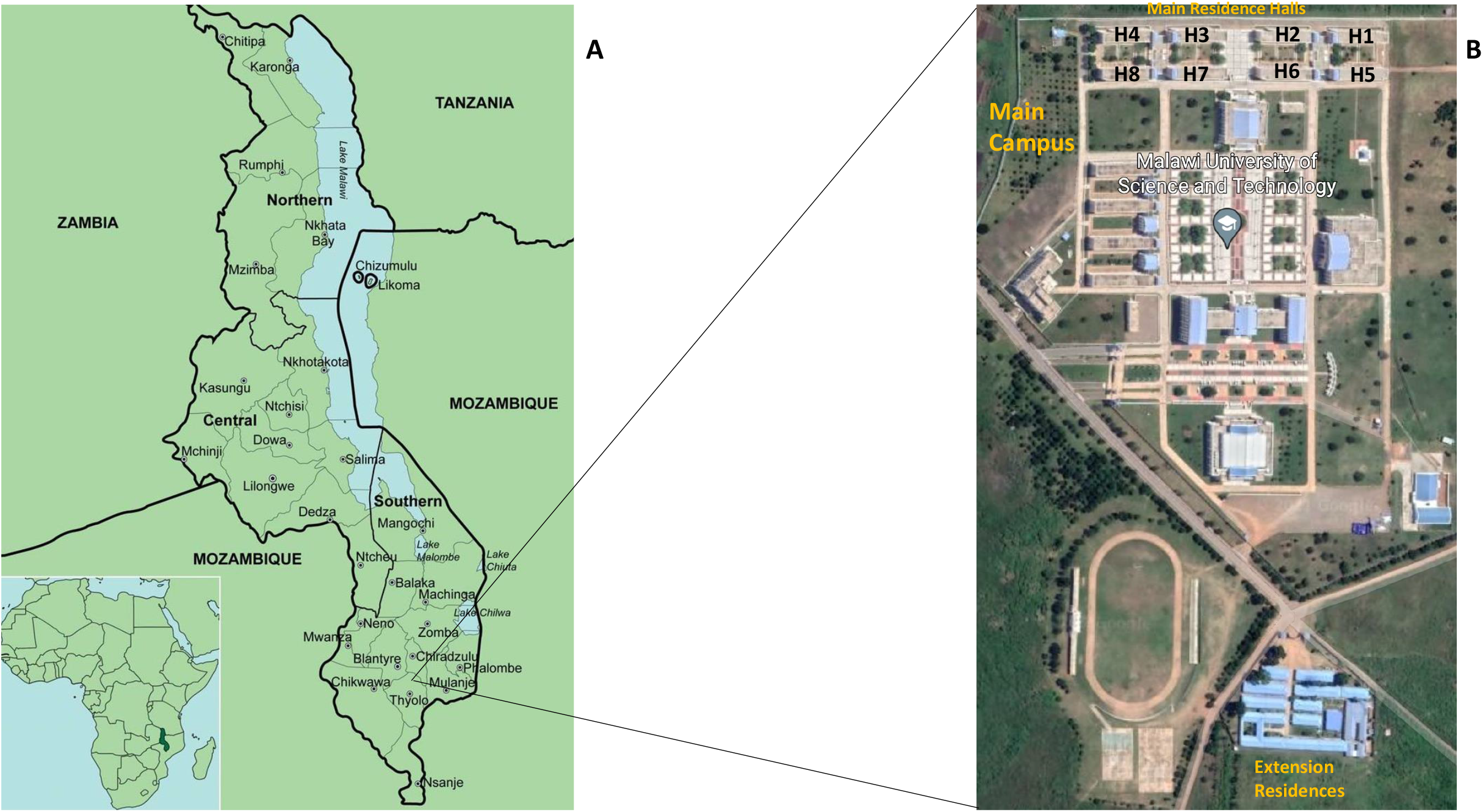
Map of the geographical location of MUST (A) showing the location of Malawi in Africa and the location of MUST in Thyolo district in southern Malawi. Also shown is a detailed campus map (B) showing the main campus and halls of residence (H1 to H8) at the top of the picture as well as the extension residences at the bottom right hand corner.

The total campus student population consists of 2530 students enrolled in 20 undergraduate programs. There are 50 students enrolled in 2 postgraduate programs who are not part of the normal campus population as their programs are conducted on a block-release basis. There is limited off campus accommodation available in the immediate community and therefore all undergraduate students are accommodated in student housing on campus.

### Preparation and resumption of in-person classes

After having moved completely to online modes of delivery, it was announced that there would be a return to in-person learning commencing in March 2021. In preparation, the MUST COVID-19 Taskforce undertook a series of measures to ensure infection prevention on campus. These included modification of spaces through posting of signage; placement of hand-wash stations at various locations on campus; development of standard operating procedures (SOP’s) and plans for transportation, sporting and other activities; modification of the student rules and regulations to reflect COVID prevention measures; establishment of a SARS-CoV-2 testing facility and a testing team; SOP’s for referral of students and staff with respiratory and other potentially COVID related symptoms; development of a communication policy and the establishment of an isolation facility.

A strategy was agreed to decongest the campus by splitting the students into two groups. Group 1: which comprised first year and final year students (March to June) n = 1100 and Group 2: which had second year and third year students (June to August) n = 1450

### Routine testing specimen collection

Individuals presenting at the MUST Clinic with any respiratory symptoms, fever or any other potentially COVID-19 related symptoms were referred to the MUST SARS-CoV-2 Molecular Laboratory for testing. All persons being tested were assisted by a trained professional in completing a case-based reporting form provided by Public Health Institute of Malawi (PHIM). Nasopharyngeal swab specimens were collected by trained personnel and placed in a 3-mL tube of viral transport medium (Mantacc MBT-010, Miraclean Technology, Shenzhen, China) to be used for both antigen testing and RT-PCR testing.

### RT-PCR on nasopharyngeal swab specimens

Viral RNA was extracted from 200 ul of viral transport media using QIAmp Viral RNA Mini Kit (Qiagen, Maryland, USA) following manufacturer’s protocol. RT-PCR was carried out using the 1-Step RT-PCR Polymerase mix with LightMix® SarbecoV E-gene and Modular SARS-CoV-2 RdRP primer-probe kits (TIB MOLBIOL, Berlin Germany).

Specimens were determined to be positive for SARS-CoV-2 when the E gene and RdRP genes were detected with an exponential growth curve and a cycle threshold (Ct) value <31, and negative when these genes could not be detected. The test was considered positive even if one gene was detected. The quality of the nasopharyngeal specimen extracted was determined by analyzing the curve generated with the human *rnaseP* housekeeping gene.

### Mass testing

Students, academic staff, administrative staff, technical and support staff were requested to present themselves for mass testing at the MUST Clinic. Testing was voluntary and all members were advised through e-mail about the importance of testing in understanding the scale of the outbreak and achieving an effective response that would allow the resumption of normal activities. Nasopharyngeal swabs were collected at MUST by clinical and laboratory staff from MUST and Blantyre District Health Office. Samples were sorted and processed at MUST using the method described above, or were transported to Blantyre DREAM Laboratories and the qRT-PCR assay was performed per the Abbott RealTi*m*e SARS-CoV-2 assay EUA product insert.

### Contact tracing

All persons who tested positive for SARS-CoV-2 were notified by phone using contact number entered on the case-based surveillance form. Individuals were asked to provide the names and contact details of their roommate or roommates as well as their 5 closest contacts over the preceding five days. Nasopharyngeal swabs were taken from the contacts within 24 hours or on the same day where possible for RT-PCR. Contact tracing was continued throughout the mass testing exercise.

### Isolation and discharge

Isolation was carried out within 24 hours of obtaining a positive PCR result. Students were escorted to the facility by a team from the Directorate of Student Affairs (DSA) during early evening hours. Meals were provided initially through arrangement for purchase and delivery from service providers close to campus. MUST management then stepped in to provide funding for meals for all isolated students from the 10^th^ of the outbreak until the isolation facility was cleared. Students were isolated for a minimum of 10 days if their PCR returned a CT value > 25. Students with CT values ≤ 24 were kept in isolation for a minimum of 14 days. Students isolating for 10 days were subjected to a follow-up PCR before release while those isolating for 14 days were given a rapid antigen test and examined by clinical officer if they had experienced symptoms during the second week of isolation. Those that had remained asymptomatic for the duration of their isolation were released without a follow up test after 14 days.

### Sequencing

Samples were shipped to the KwaZulu-Natal Research Innovation and Sequencing Platform’s (KRISP) laboratory in Durban, South Africa for sequencing. RNA extraction was carried out on the automated Chemagic 360 instrument using the CMG-1049 kit (Perkin Elmer, Hamburg, Germany) according to the manufacturer’s instructions. Complementary DNA (cDNA) synthesis was performed using random hexamer primers from the SuperScript IV reverse transcriptase synthesis kit (Life Technologies), followed by multiplex PCR according to the nCoV-2019 ARTIC network sequencing protocol (https://artic.network/ncov-2019). AmpureXP purification beads (Beckman Coulter, High Wycombe, UK) were used to purify the PCR products in a 1:1 ratio and further quantified on the Qubit 4.0 instrument (Life Technologies Carlsbad, CA) using the Qubit dsDNA High Sensitivity assay according to the manufacturer’s instructions.

### Library Preparation and Illumina Sequencing

Library preparation was done using the Nextera Flex DNA library preparation kit and Nextera CD indexes (Illumina, San Diego, CA, USA) according to manufacturer’s instructions. The libraries were quantified using the Qubit 4.0 instrument (Life Technologies) and the Qubit dsDNA High Sensitivity assay kit. Each sample library was normalized, pooled and denatured. Libraries were sequenced on the Illumina MiSeq platform using a MiSeq Reagent Kit v2 (500 cycles).

### Sequence analysis

Sequenced genomes from the outbreak were assembled using Genome Detective 1.132 (8) and mapped to the Wuhan-Hu-1 SARS-CoV-2 reference. The resulting assemblies where cleaned in Geneious. MUST sequences were analysed against a global representative set of sequences from around the world including all Malawian SARS-CoV-2 sequences contained within the GISAID sequence repository (date of access 1^st^ July 2021) using the COVID-19 NextStrain build platform (https://github.com/nextstrain/ncov).

## Results

### Epidemic Timeline and description of outbreak

The outbreak began with two confirmed cases referred from the clinic with COVID related symptoms (Figure 2). Both individuals were requested to isolate at home or transfer to the isolation facility that had been set-up at MUST within 24 hours. Immediate contacts of both cases traced and requested to undergo testing the following day. All classes and cohorts that had contact these two cases were immediately suspended and all lecturers were notified. Targeted testing on day-2 yielded nine cases from 42 tests.

**Figure 2:**
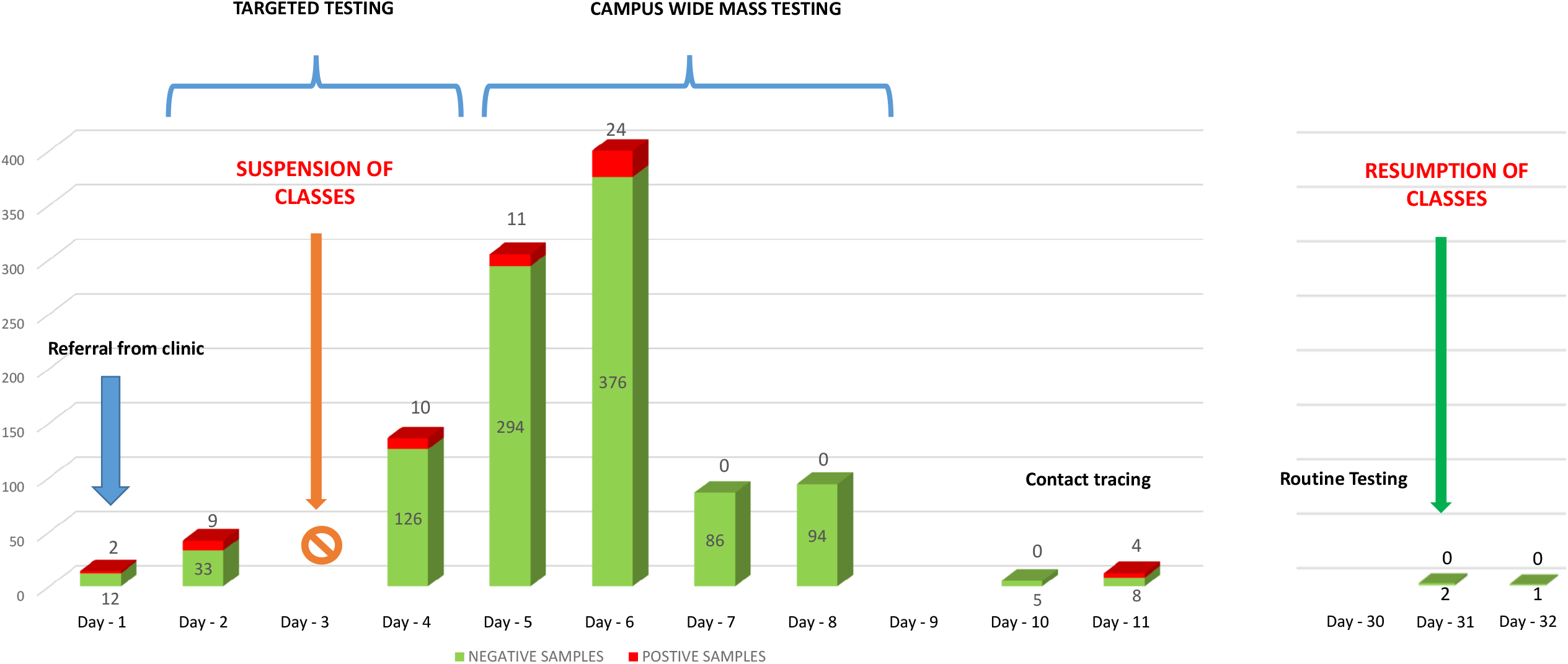
Timeline of cases at MUST during the months of April and May 2021. Dates are placed along the bottom axis with a 17 day gap between the 17^th^ April and the 3^rd^ May where no positive test result was obtained. Positive test results shown in red and negative tests shown in green. The numbers of tests are indicated on or close to each relevant bar. The suspension of classes on the 8^th^ of April and resumption of classes on the 5^th^ of May are indicated as are the 4 days of the campus-wide mass testing exercise on the 10^th^, 11^th^, 12^th^ and 13^th^ of April.

Having identified 11 cases out of 54 tests (20% positivity rate) in two days of testing it became apparent that there was a significant cluster of infections among students on campus. All in-person learning activities on campus were halted and teaching moved to online modes of delivery. An initial suspension of 21 days was announced, and the situation would be reviewed before resumption of classes.

Mass testing for all staff and students was arranged beginning on day 4. Given the limited capacity of the MUST laboratory, priority was given to testing groups (study programs or residences) that had epidemiological links to the cases found thus far. A total of 136 tests were conducted with 10 positive results.

On day 5, additional testing capacity was brought on board with the assistance of the Blantyre District Health Office and the Blantyre DREAM Molecular Laboratory. This allowed us to conduct 305 tests on day 5 and 400 tests on the 6. These returned 11 and 24 positive results, respectively. A total of 86 and 94 tests were conducted on day 7 and day 8 and all were negative. No tests were conducted on day 9 and the mass testing exercise was completed.Subsequently, we reverted to testing symptomatic individuals referred from the clinic. Five negative tests were done on day 10. Some referrals and contact tracing on day 11 resulted in 4 new cases from 12 tests conducted.

There were no more referrals from the clinic for the next 20 days and all tests conducted were follow-up tests of isolated individuals seeking discharge to and a return to their campus accommodation.

### Coverage of staff and students by mass testing

The mass testing exercise was open to everyone at MUST and although it was not actively encouraged, community members from surrounding areas were also allowed to test. A total of 1,094 tests were conducted in the month of during the ourbreak.

Staff were categorised in two groups, namely academic and support staff as well as outsourced service staff. Outsourced service staff consisted of those in catering services, cleaning services, landscaping services and security services. In total, 161 (40%) staff members out of 404 were tested (Figure 3). Only 6.3% of academic and support staff tested while 87.4% of outsourced service staff tested.

**Figure 3:**
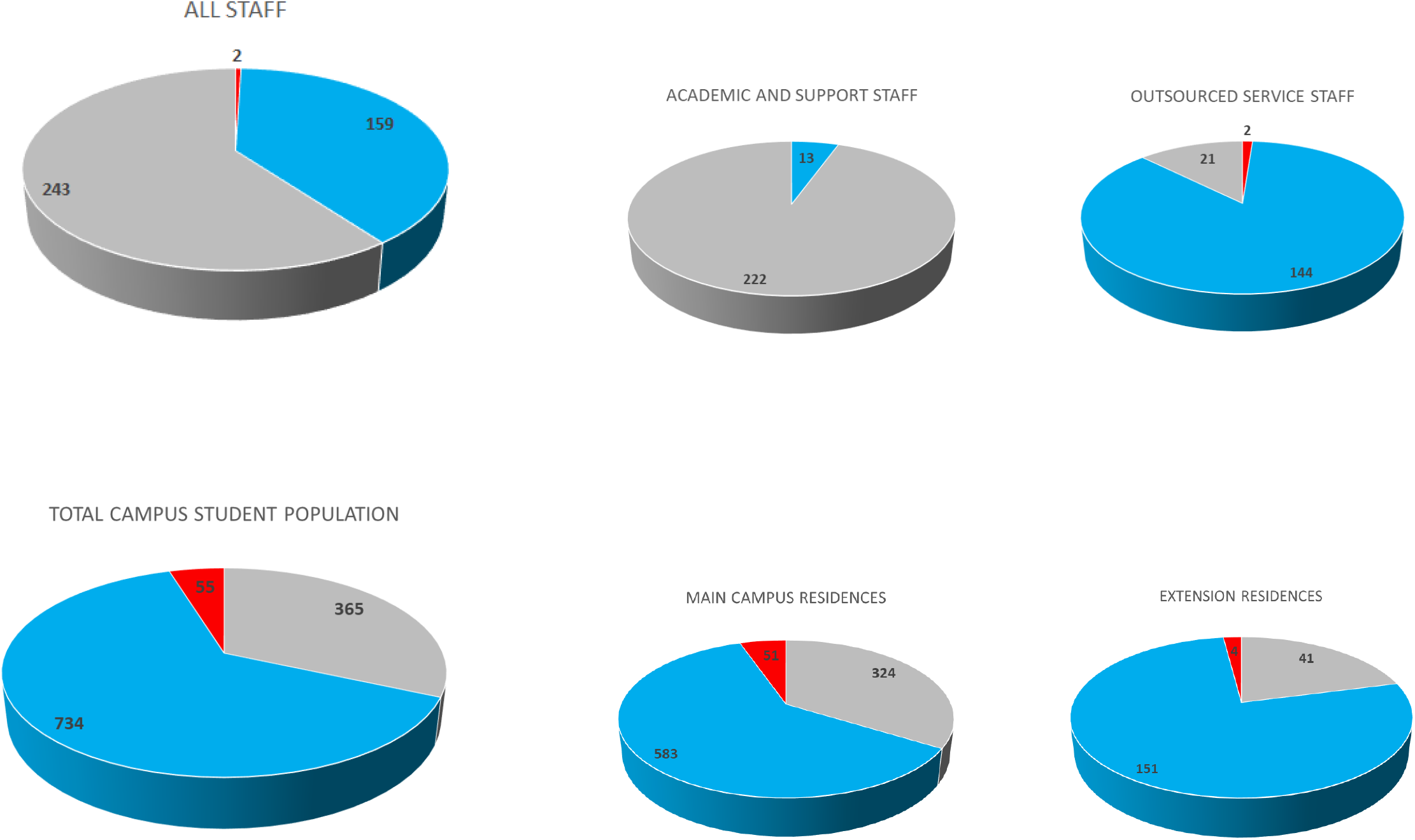
Pie charts illustrating summaries of cases among staff and students from Table 1: Red sections indicate positive cases, blue sections indicate negative tests and grey sections indicate untested individuals. Numbers of positive cases are indicated in the respective sections. Staff are grouped into two; academic and support staff as well as outsourced service staff. Students are grouped into those residing on the main campus and those residing in the extension campus.

A total of 789 tests were conducted among students from a population of 1154 representing 68.4% of students (Figure 3). There were 55 positive tests giving a total positivity rate of 6.97%. A total of 51 out of the 55 (92.7%) positive cases among students were housed on the main campus where most students reside putting the positivity rate there at 8.04%. This was significantly higher (χ^2^ (1, 5.733), p=0.0166) than the extension camp which had a positivity rate of 2.58% among its population of 196 students.

**Table 1A:** Summary statistics for PCR testing of MUST Staff during the month of April 2021.

**STAFF SUMMARY TABLE**
| Variable | TOTAL | PCR negative | PCR positive | Not Tested |
| --- | --- | --- | --- | --- |
| **Sex** |  |  |  |  |
| Female | 136 | 60 | 1 | 76 |
| Male | 268 | 105 | 1 | 163 |
| **Population** |  |  |  |  |
| Academic and Support Staff | 235 | 13 | 0 | 222 |
| Outsourced Service Staff | 169 | 144 | 2 | 21 |
| **Clinical status** |  |  |  |  |
| Asymptomatic | 152 | 150 | 2 |  |
| Symptomatic | 15 | 15 | 0 |  |

**Table 1B:** Summary statistics PCR testing of students at MUST during the month of April 2021.

**STUDENT SUMMARY TABLE**
| Variable | TOTAL | PCR negative | PCR positive | Not Tested |
| --- | --- | --- | --- | --- |
| **Sex** |  |  |  |  |
| Female | 292 | 183 | 12 | 97 |
| Male | 860 | 517 | 43 | 300 |
| **Population** |  |  |  |  |
| First Year Students | 592 | 449 | 14 | 129 |
| Final Year Students | 560 | 251 | 41 | 268 |
| **Clinical status** |  |  |  |  |
| Asymptomatic | 669 | 637 | 32 |  |
| Symptomatic | 86 | 63 | 23 |  |

The testing coverage among students was significantly lower (χ^2^ (1, 12.89), p=0.0003) at 65% on the main campus compared to 74% at the extension camp.

There were 146 tests which were unidentifiable as students or staff members. Among these, there were no cases detected.

### Distribution of cases on campus

An analysis was done of the location of each individual case in the main campus residences by their room allocation (figure 4). In one residence 24% of occupants tested positive while all other residences had a positivity rate below 5%. In each case identified in the residences the roommate was treated as a contact and was encouraged to present for testing.

**Figure 4:**
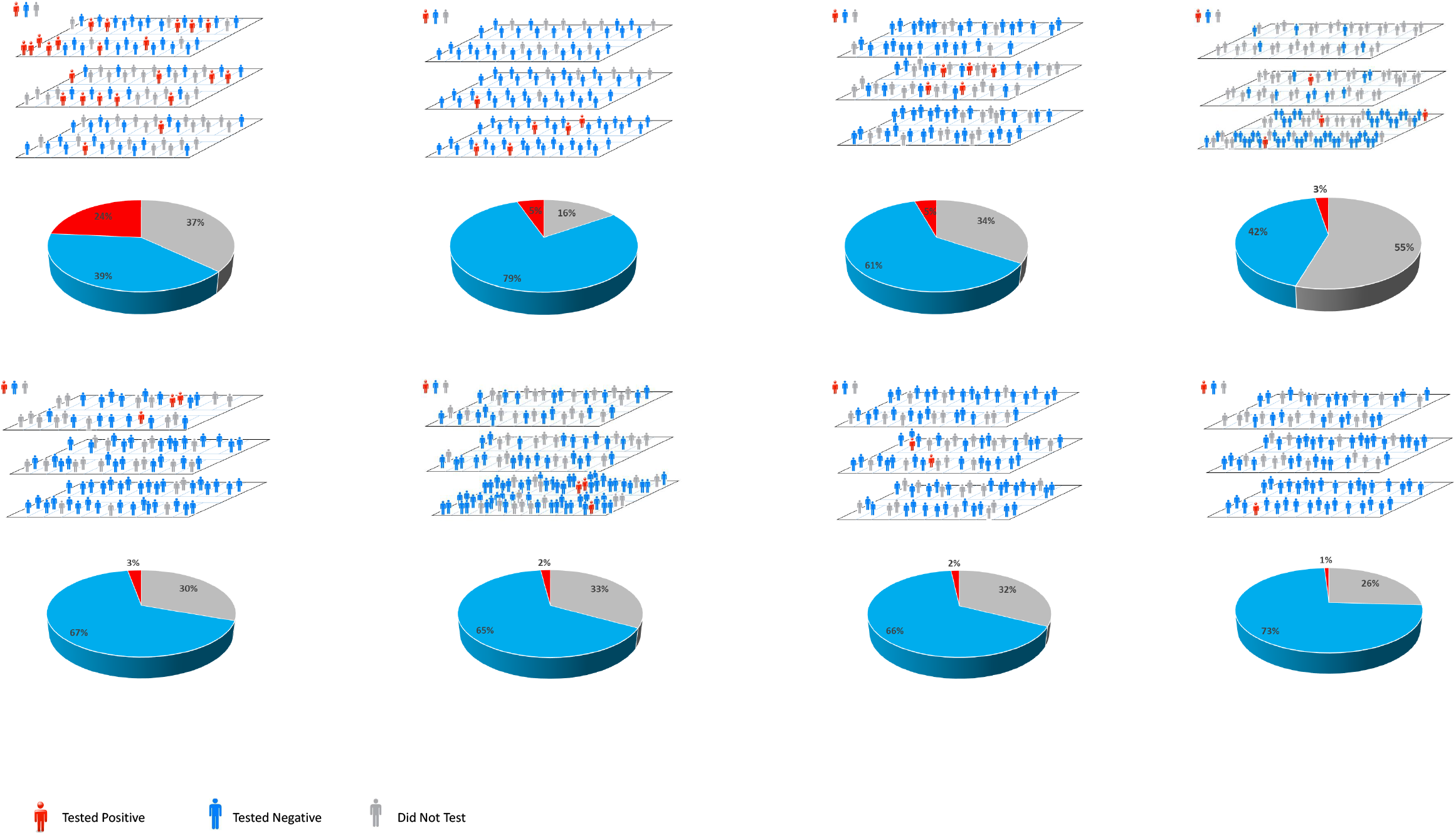
Graphical display of distribution of cases in halls of residence on the main campus accompanied by pie charts showing percentages of infected in residences. The residences are displayed in order of highest incidence to lowest (and therefore randomised). Positive cases are indicated in red, those testing negative are in blue while those that did not get tested are shown in grey.

### Testing coverage among students

Some trends were evident in the willingness of different groups of students to test as evidenced by the percentages of those who submitted biological samples. Taken overall there was no significant gender difference in positivity rate and testing coverage among students. Male students had a positivity rate of 7.7% and achieved 65.12% testing coverage while female students had a positivity rate of 6.15% and achieved 66.78% testing coverage. However, there were no female students in the extension camp and therefore the gender differences were re-examined only among students in the main campus. No significant differences were observed in test positivity however male students did have a significantly reduced level of testing coverage of 61.0% (χ^2^ (1, 6.865), p=0.00879).

There were significant differences observed in test positivity and testing coverage between the first year (n=592) and final year students (n=560) (Year 4 or year 5 students depending on the program of study). There was a lower test positivity of 3.02% among first year students compared to 14.04% positivity rate among final year students (χ^2^ (1, 86.575), p<0.00001). First year students had higher testing coverage of 78.2% compared to 52.14% among final year students (χ^2^ (1, 32.18), p<0.00001).

The same trend was observed on detailed analysis residences which had mixed populations of freshmen and final year students. In one case, the combined testing rate was 58.2%.

However, when disaggregated, testing coverage for first year students was 69.4% while that for final year students was only 39.5% of students tested (χ^2^ (1, 33.071), p<0.00001). In a second case, the combined testing coverage was 66.4% but was disaggregated as 80.3% of first year students and 52.5% of final year students tested (χ^2^ (1, 11.8197), p=0.000586). The highest coverage of 85% was achieved in a residence which was all female and first year.

### Virus sequencing

A total of 38 samples were successfully sequenced. All were found to be Nextstrain clade 501.Y.V2 that corresponds to Pango lineage B.1.351 or WHO label Beta variant of concern (VOC). Phylogenetic analysis of the sequenced specimens from MUST against other Malawian and global sequences revealed that there were multiple introductions of the virus on the MUST campus, including 4 that lead to the April outbreak (Fig 5A) (https://nextstrain.org/groups/ngs-sa/COVID19-MW-MUST-2021.07.07?c=outbreak&f_division=Malawi). While the sequences from January and February were sampled from COVID positive staff present on campus in the absence of students and were assigned to the Beta variant, it is clear from phylogenetic analysis that none of these were the source of the student outbreak in April (Fig 5B). Most of the sequences from the April outbreak belong to two distinct transmission clusters within the Beta clade, the largest one containing at least 25 staff/students from MUST (Fig 5D) and the other one containing at least 6 individuals (Fig 5C). Two cases of the April outbreak did not go beyond a single case.

**Figure 5:**
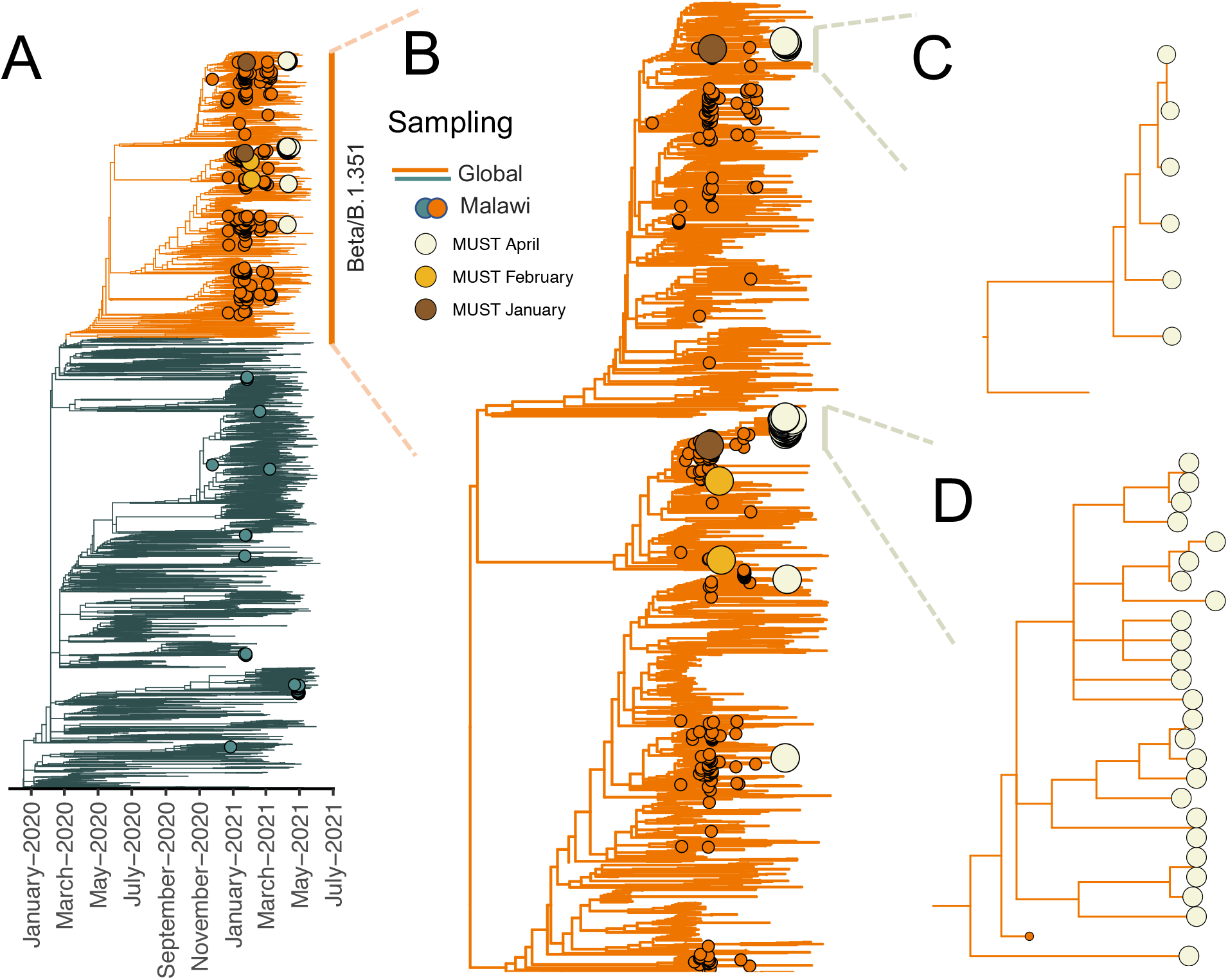
Phylogenetic analysis of the MUST outbreak. A) Time-scaled Maximum-Likelihood (ML) tree containing 336 Malawian sequences publicly available on GISAID, 38 of which were sampled from MUST, along with a representative set of 4648 other global sequences. The Beta/B.1.351 clade is annotated in orange while other clades are annotated in grey. Malawian genomes are indicated with tip points matching the clade annotation and MUST sequences are annotated with other distinct colours depending on the time of sampling (off-white, yellow, brown). B) Zoomed-in view of the Beta clade showing multiple introductions of the virus into the MUST campus, with two large transmission clusters from the April outbreak indicated. C) Transmission cluster containing 6 specimens from MUST. D) Transmission clusters containing 25 specimens from MUST and one other Malawian genome.

### Resumption of in-person classes

On day 31, after having 18 days without recording a new case, in-person classes were resumed. By day 33 all individuals who had been kept in the isolation facilities had recovered and been discharged.

## Discussion

Societies and institutions are still grappling with how to effectively respond to and manage successive waves of highly disruptive COVID infections and outbreaks. The semi-contained university setting provides a level of administrative control that is often unavailable in the general population. A combination of vaccination and non-pharmaceutical interventions need to be deployed in an effective response. However, Malawi remains one of the least vaccinated countries and Africa the least vaccinated continent highlighting the importance of non-pharmaceutical interventions. The currently used TTI strategies are likely to remain a critical part of outbreak management and are important for making students and staff feel safe and enable the rapid return to normal activities. These strategies have to overcome several challenges to be effectively implemented. These include the presence of asymptomatic yet infectious carriers (9,10), opposition and reluctance to testing (11) and capacity limits of testing as well as contact tracing (6).

The experience at MUST shows that challenges exist in convincing all staff and students to present themselves for testing. Turnout was especially low for academic staff, and this was likely due to the fact that almost all of them reside off campus in Blantyre or Zomba several kilometres away. Support staff are mainly resident in the surrounding communities and showed a greater willingness to test accompanied with a very low incidence. Related to this was the unexpectedly large turnout of community members (n=146) who took advantage of the mass testing exercise to know their COVID-19 status and the complete absence of cases among this group. This trend is reflective of the reported lower seroprevalence in rural Malawians seen in the second wave. However, this pattern is beginning to shift with studies reporting higher seroprevalence among rural communities in the more recent delta driven wave (M Crampin, personal communication, July 13, 2021). This further highlights the importance of controlling on-campus transmission as a means of protecting the adjacent community which often has less access to testing and care than urban communities do. Our findings also suggest that the community surrounding the campus would benefit from and perhaps welcome access to COVID testing through MUST.

Among students, gender differences were observed in the percentage of male and female students presenting for testing with female students being more cooperative with the mass testing exercise. The difference observed in this study is reflective of similar trends seen in several multicultural studies of university students’ health seeking behaviour and outcomes (12) and even health literacy (13). Research conducted in Malawi shows that males spend almost no time seeking healthcare (14) and that this is associated with feelings of diminished masculinity (15,16). Health interventions will likely be more effective if they give particular focus and attention to male students (12).

An important finding in this study was that senior students (year 4 and 5) had higher COVID-19 incidence but fewer of them presented for testing compared to freshmen. The reasons for senior students being less cooperative and compliant to preventive measures is an important area of research that needs to be explored to better inform interventions at universities and schools. Refusal to test presents a danger to members of any community particularly in the case of SARS-CoV-2 which is a serious communicable disease. Ensuring that the importance of testing and the consequences of refusal are well articulated and understood by all is an important part of any strategy. Lessons can be drawn from other diseases and continued development of the ethical and legal implications of refusal to test will remain an important public health policy issue.

Further studies are required to better understand other barriers to testing. Anecdotally, several students expressed that the use of nasopharyngeal swabs is a major discouraging factor and staff and students welcomed testing with anterior nasal swabs where these were available.More work is also required to understand the experience and psychosocial impact of being moved to isolation facilities. Provision of food to students in isolation for example, was viewed as a strong incentive by some students.

Response to a campus outbreak requires effective planning, commitment of resources and focussed implementation and coordination between campus health services, laboratory services, student affairs directorate and the registry. It is important also that universities work closely with local health authorities to effectively protect the local communities and report case numbers and surveillance data to the national response coordinators. This study reports an outbreak driven by multiple introductions of the beta variant. As more transmissible variants such a delta emerge and drive the next waves of infection, campus response teams will need to act in a more coordinated, decisive and aggressive manner to control any future outbreaks and minimise the disruptive effect of COVID.

## Data Availability

All data produced in the present study are available upon reasonable request to the authors

## Acknowledgements

We are grateful to Seeding Labs (Boston, MA, USA) for their kind donation of equipment and to the Ministry of Health in Malawi for facilitating the establishment of the COVID Molecular Testing Laboratory at MUST and for support with training, reagents and data management. We appreciate the technical assistance of Watipaso Kasambara and her team especially the late Aisha Senga for her assistance. We are thankful to Dr Kondwani Jambo and Dr Sara Suliman for the critical discussion and sharing of ideas. We appreciate the MUST COVID Taskforce and the assistance of Blantyre and Thyolo District Health Offices in the management of the response. Funding was provided by MUST management. All authors had full access to all data in the study, and the corresponding author had final responsibility for the decision to submit for publication. GB led the study team, conceptualised and drafted the manuscript. AM provided overall guidance and helped conceptualised the study. PC and TN provided critical feedback in manuscript writing. YK, JM and MK conducted laboratory assays and data processing. PK developed the tables and figures. MM and YN led the sample and data collection. MM, CM, SK and MS managed isolation and management of students. JBB and TN facilitated establishment of testing capacity and provided feedback on the manuscript. RL provided mass testing capacity. BM facilitated sequencing with the UKZN team. JG, SP, YN, UR, JES, HT, EW and TdO conducted virus sequencing and developed figures. AM

## Author Bio

Gama Bandawe (PhD) is a medical virologist and is currently a senior lecturer and head of the Department of Biological Sciences in the Academy of Medical Sciences at the Malawi University of Science and Technology (MUST). His research interests include immune responses to HIV and HIV diversity as well as emerging virus infections and public health in Malawi.

